# Developing a model for predicting impairing physical symptoms in children 3 months after a SARS-CoV-2 PCR-test: The CLoCk Study

**DOI:** 10.1101/2022.04.01.22273117

**Authors:** Manjula D Nugawela, Terence Stephenson, Roz Shafran, Bianca L De Stavola, Shamez N Ladhani, Ruth Simmons, Kelsey McOwatt, Natalia Rojas, Emily Y Cheung, Tamsin Ford, Isobel Heyman, Esther Crawley, Snehal M Pinto Pereira

## Abstract

**Importance:** Predictive models can help identify SARS-CoV-2 patients at greatest risk of post-COVID sequelae and direct them towards appropriate care.

**Objective:** To develop and internally validate a model to predict children and young people most likely to experience at least one impairing physical symptom 3 months after a SARS-CoV-2 PCR-test and to determine whether the impact of these predictors differed by SARS-CoV-2 infection status.

**Design:** Potential pre-specified predictors included: SARS-CoV-2 status, sex, age, ethnicity, deprivation, quality of life/functioning (5 EQ-5D-Y items), physical and mental health, and loneliness (all prior to SARS-CoV-2 testing), and number of physical symptoms at testing. Logistic regression was used to develop the model. Model performance was assessed using calibration and discrimination measures; internal validation was performed via bootstrapping; the final model was adjusted for overfitting.

**Setting:** National cohort study of SARS-CoV-2 PCR-positive and PCR-negative participants matched according to age, sex, and geographical area.

**Participants:** Children and young people aged 11-17 years who were tested for SARS-CoV-2 infection in England, January to March 2021.

**Main outcome measure:** one or more physical symptom 3 months after initial PCR-testing which affected physical, mental or social well-being and interfered with daily living.

**Results:** A total of 50,836 children and young people were approached; 7,096 (3,227 test-positives, 3,869 test-negatives) who completed a questionnaire 3 months after their PCR-test were included. 39.6% (1,279/3,227) of SAR-CoV-2 PCR-positives and 30.6% (1,184/3,869) of SAR-CoV-2 PCR-negatives had at least one impairing physical symptom 3 months post-test. The final model contained predictors: SARS-COV-2 status, number of symptoms at testing, sex, age, ethnicity, self-rated physical and mental health, feelings of loneliness and four EQ-5D-Y items before testing. Internal validation showed minimal overfitting with excellent calibration and discrimination measures (optimism adjusted calibration slope:0.97527; C-statistic:0.83640).

**Conclusions and relevance:** We developed a risk prediction equation to identify those most at risk of experiencing at least one impairing physical symptom 3 months after a SARS-CoV-2 PCR-test which could serve as a useful triage and management tool for children and young people during the ongoing pandemic. External validation is required before large-scale implementation.

**Key Points:** *Question:* Which children have impairing physical symptoms during the COVID-19 pandemic?

*Findings:* Using data from a large national matched cohort study in children and young people (CYP) aged 11-17 years (N=7,096), we developed a prediction model for experiencing at least one impairing physical symptom 3 months after testing for SARS-COV-2. Our model had excellent predictive ability, calibration and discrimination; we used it to produce a risk estimation calculator.

*Meaning:* Our developed risk calculator could serve as a useful tool in the early identification and management of CYP at risk of persisting physical symptoms in the context of the COVID-19 pandemic.

## Introduction

Children and young people (CYP) testing positive for SARS-COV-2 are usually asymptomatic or have a low symptom burden at the time of infection compared to adults.^1,2^ Recent studies on post-COVID sequelae (also known as ‘long COVID’), however, have shown some adults and children can have persistent symptoms for months after acute infection.^3,4^ A recent systematic review of persistent symptoms following SARS-COV-2 infection, found most reported persistent symptoms were no more common in SARS-COV-2 positive than in SARS-COV-2 negative CYP, with only small increases in cognitive difficulties, headache, loss of smell, sore throat and eyes.^5^

Similar to the successful use of predictive models for cardiovascular disease e.g., in the UK^6^ and the US^7^, predictive models can help identify CYP at highest risk of experiencing persistent symptoms and direct them towards relevant care. This is particularly important during the pandemic when health services are under increased pressure.^8^ A systematic review identified over 100 diagnostic and prognostic models for SARS-COV-2, mainly relating to acute outcomes e.g., mortality, ICU admission and length of hospital stay.^9^ With the exception of two studies, however, most were considered low quality due to non-representative selection of controls, inadequate exclusions, high risk of model overfitting and unclear reporting.^9^ Based on predictive model quality assessment tools^10^ and model development guidelines,^11^ the two models mentioned above (the Jehi diagnostic model^12^ and 4C mortality score^13^) and a third model (QCOVID^14^) are considered as higher quality predictive models for SARS-COV-2 because of large sample sizes,^15^ appropriate modelling techniques^16^ and suitable internal validation and reporting.^11^ Of these three models, the 4C and QCOVID models were developed in adult populations (age≥18 years) whereas the Jehi model was developed in all patients who were tested for SARS-CoV-2 at all Cleveland Clinic locations in Ohio and Florida, USA, regardless of age and included 11,672 patients (median age: 46.89 years among SARS-COV-2 negatives; 54.23 years among SARS-COV-2 positives).

There are very few predictive models for the potential long-term effects of SARS-COV-2 infection, and those that exist have focused mostly on adults. Sudre and colleagues, focused on identifying the characteristics and predictors of post-COVID sequelae in a sample of 4,182 adults who reported testing positive for SARS-CoV-2 and found those experiencing more than five symptoms during the first week of illness were more likely to report ‘long COVID’.^17^ Recent large national cohort studies of CYP are consistent with the recent systematic review^5^ and have found little difference in ‘long COVID’ symptom prevalence between SARS-CoV-2 positive and SARS-CoV-2 negative CYP.^4,18^ As acute SARS-CoV-2 infection remains predominantly a mild infection in CYP and the cumulative incidence of infection increases, the incidence of post-COVID sequelae and the extent to which it is distinct from pandemic-related symptoms resulting from national lockdowns, school closures and social isolation, is a critical factor in health policy decisions. We, therefore, aimed to develop and internally validate a prediction model for experiencing at least one impairing physical symptom in CYP 3 months after a PCR-test and to determine whether the impact of these predictors differed by SARS-CoV-2 infection status. The outcome examined here aligns with our previously described Delphi definition of post-COVID sequelae.^19^

## Methods

We use data from the CLoCk study: a national cohort study of SARS-CoV-2 PCR-positive CYP aged 11-17 years living in England who were matched at study invitation, on month of test, age, sex, and geographical area, to SARS-CoV-2 test-negative CYP selected from the national testing database at Public Health England (now UKHSA).^20^ Test-negative CYP who self-reported subsequently having a positive SARS-COV-2 PCR-test were excluded.^4^

Here we examine a previously described study subset.^4^ Briefly, from a total of 50,836 CYP who were approached, 7,096 (3,227 SARS-COV-2 positive, 3,869 SARS-COV-2 negative) who completed the CLoCk questionnaire sent to them 3 months after their PCR test during January-March 2021 (median time between testing and questionnaire: 14·9 weeks [25^th^,50^th^ centiles: 13·1,18·9]) were included. The questionnaire included demographic characteristics, elements of the International Severe Acute Respiratory and emerging Infection Consortium (ISARIC) Paediatric COVID-19 follow-up questionnaire,^21^ and the recent Mental Health of Children and Young people in England surveys.^22^ CYP responded to 21 questions on physical symptoms at time of testing (e.g., cough, tiredness, etc.). They rated their general physical and mental health before SARS-CoV-2 testing in two separate questions using a five-category Likert scale. The prevalence of ‘very poor’ was low; for analysis, we recoded these variables into four categories (very poor/poor to very good). Quality of life/functioning before testing was measured via the EQ-5D-Y scale,^23^ and feelings of loneliness by the UCLA Loneliness scale.^24^ The Index of Multiple Deprivation (IMD), was calculated from the CYP’s small local area level based geographic hierarchy (lower super output area) at the time of the questionnaire and used as a proxy for socio-economic status. We examine IMD quintiles from most (quintile 1) to least (quintile 5) deprived (Table 1).

**Table 1.**
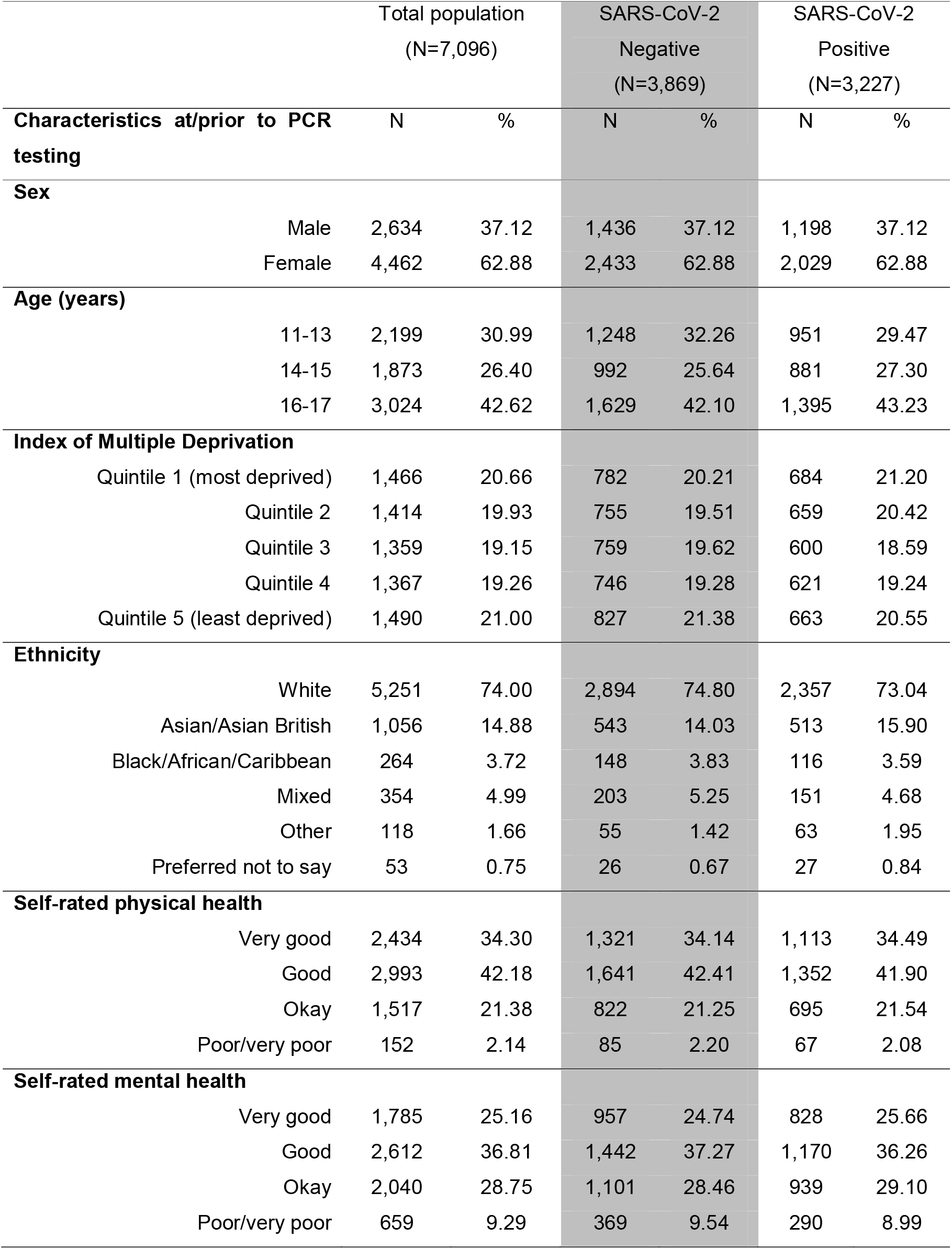
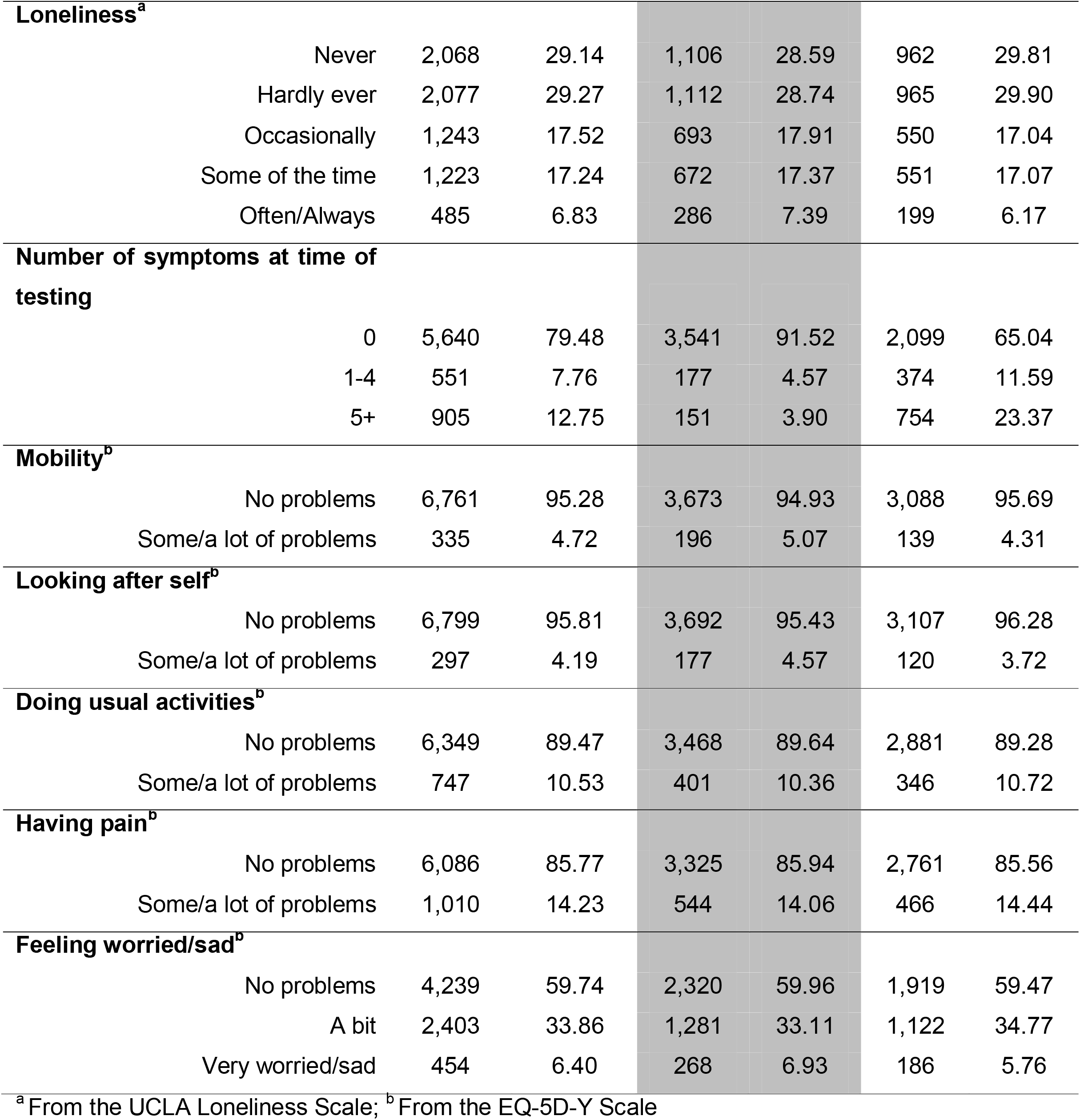
Baseline characteristics (frequencies and percentages) of participants who completed the 3-month questionnaire, overall and stratified by SARS-CoV-2 status

## Outcome

### experiencing at least one impairing physical symptom

The outcome was defined as having one or more physical symptom for at least 12 weeks after initial testing even if symptoms waxed and waned over that period, and the symptoms affected their physical, mental or social-well-being while interfering with some aspect of daily living (e.g., school, home, relationships). This outcome aligns with our previously described Delphi definition of post-COVID sequelae.^19^ Using data from the questionnaire on physical symptoms and the EQ-5D-Y scale at the time of the questionnaire (i.e. approximately 3 months after the PCR-test), we operationalized our outcome as having at least 1 physical symptom and experiencing at least some problems on any one of the five EQ-5D-Y questions (Table 2).

**Table 2.**
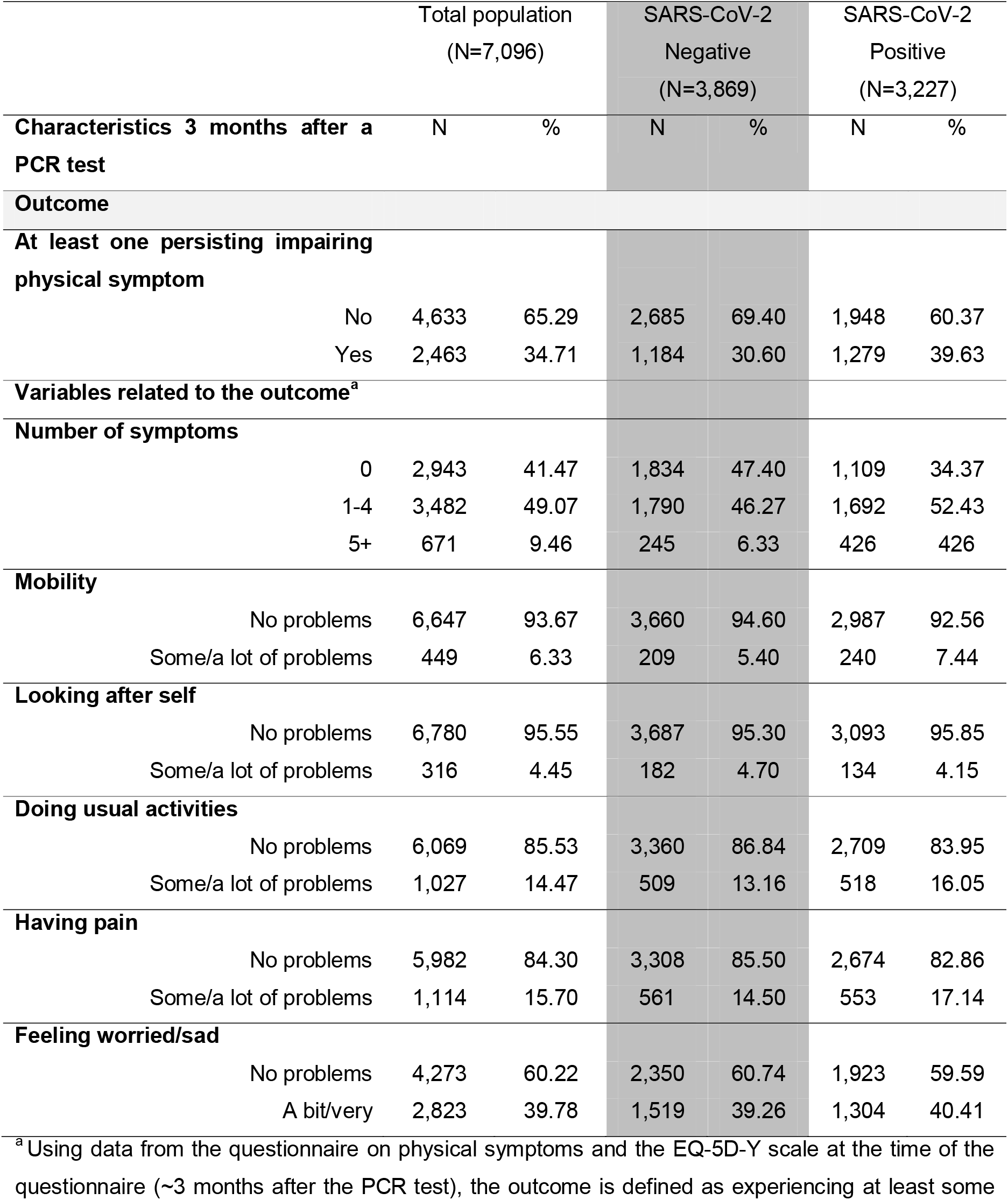
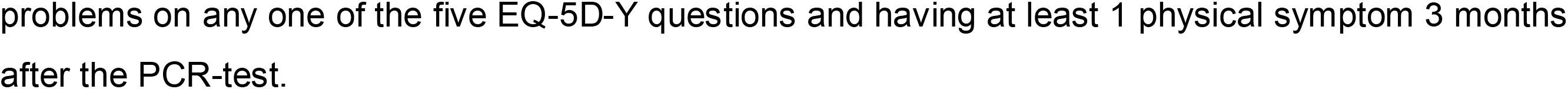
Prevalence (frequencies and percentages) of at least one persisting impairing physical symptom 3 months after a PCR test and related variables^a^, overall and stratified by SARS-CoV-2 status

### Potential predictors

Pre-specified potential predictors were chosen based their distribution in the dataset and their association with the outcome. In addition to SARS-COV-2 status, we considered 13 predictors including demographics (sex, age, ethnicity and IMD), prior quality of life/functioning (assessed by 5 items from the EQ-5D-Y scale), prior physical and mental health and feelings of loneliness prior to the CYP’s PCR-test. We also included number of physical symptoms experienced at testing (details in Table 1).

### Sample size and missing data

The sample size was pre-defined by study design. We, therefore, assessed whether our study was sufficiently powered to estimate the overall outcome risk, and how many predictor parameters could be considered before overfitting/precision becomes a concern.^15^ Using the pmsampsize STATA package^15^ we considered: i) small overfitting (i.e., a shrinkage factor of predictor effects ≤10%), ii) small absolute difference of 0.05 in the model’s apparent and adjusted Nagelkerke’s R-squared value, and iii) precise estimation within ±0.05 of the average outcome risk in the population. We also assumed an outcome prevalence of 34.7%, C-statistic of 0.80 and 61 parameters. Accordingly, the minimum sample size required was 1,943 (actual sample=7,096); the events-per-candidate predictor parameter value was 11.05. The dataset had no missing data.

## Statistical Analysis

We assessed the extent to which SARS-COV-2 status and our 13 potential predictors were correlated by considering pairwise Cramer’s V correlation coefficients. All potential predictors were categorical variables, with the exception of age and number of symptoms at testing. We determined the appropriate functional form for the relationship between age and the log-odds of the probability of the outcome by modelling the relationship (i) linearly, (ii) categorically (11-13, 14-15, 16-17 years), (iii) with linear and quadratic terms, and (iv) using fractional polynomials with up to 2 degrees. Similarly, we examined the most appropriate functional form for number of symptoms. The functional form with the lowest Akaike Information Criterion (i.e., the best fitting model) was used in building our prediction model.

We used logistic regression to address our aim of predicting at least one impairing physical symptom in children 3 months after their PCR-test, allowing for an interaction between each potential predictor and SARS-COV-2 status to determine whether the relationship between the potential predictor and outcome differed by SARS-COV-2 status. We first examined univariable associations between each predictor and at least one impairing physical symptom, in the total population and stratified by SARS-CoV-2 status. Next, we built a multivariable prediction model using a stepwise backward (p<0.200) and forward (p<0.157) elimination procedure.^25^ Variables included in the stepwise selection procedure included all potential predictors, SARS-COV-2 status and interaction terms between potential predictors and SARS-COV-2 status (61 potential parameters in total).

Model performance was measured using calibration and discrimination measures. Calibration, (i.e., agreement between observed and predicted probabilities of our outcome) was assessed using calibration plots, calibration-in-the-large and calibration slope statistics.^16,26^ Model discrimination, (i.e., the ability of our model to differentiate between CYP who had at least one impairing symptom 3 months post-test and those who did not) was quantified using the C-statistic (values≥0.7 indicate strong discrimination). The internal validity of our final model was assessed using 100 bootstrap samples which were drawn with replacement.^16^ We estimated the level of model overfitting (optimism) in our dataset using the bootstrap samples and adjusted for optimism using a uniform shrinkage factor (the average calibration slope from each of the bootstrap samples). The original β coefficients were multiplied by the shrinkage factor to obtain the optimism adjusted coefficients; the model intercept was re-estimated based on these shrunken model coefficients generating the final model.^11,26^

Data management and analysis was performed using STATA16. We followed guidelines by the Prognosis Research Strategy (PROGRESS)^27-30^ Group; the model development and validation phases particularly followed suggested methods. ^26,29-31^ The study is reported according to the Transparent Reporting of a multivariable prediction model for Individual Prognosis or Diagnosis (TRIPOD) statement (eTable 1).^11^ The study was approved by Yorkshire and the Humber–South Yorkshire Research Ethics Committee (reference: 21/YH/0060).

## Results

Of the 7,096 CYP (3,869 SARS-CoV-2 negative, 3,227 SARS-CoV-2 positive) in our analytic sample, 26% (1845/7096) were non-white, 62.9% (4,462/7096) were females and there were more older than younger CYP (42.6% 16-17-year-olds vs. 31.0% 11-13-year-olds) (Table 1). Three months after their PCR test, 65.6% (2,118/3,227) of SAR-CoV-2 PCR-positives had at least one physical symptom (Table 2) and 39.6% (1,279/3,227) had at least one impairing physical symptom. This compares with 52.6% (2,035/3,869) and 30.6% (1,184/3,869), respectively, in test-negative CYP.

### Univariable associations

SARS-CoV-2 status and the 13 potential predictors were not strongly correlated (Cramer’s V<0.50 for all possible pairwise correlations). IMD did not predict the outcome (Table 3). The predictive effect of feelings of loneliness, problems with mobility, doing usual activities and having pain before testing differed by SARS-CoV-2 status, with a general pattern of higher odds among test-negatives (Table 3, stratified associations).

**Table 3.**
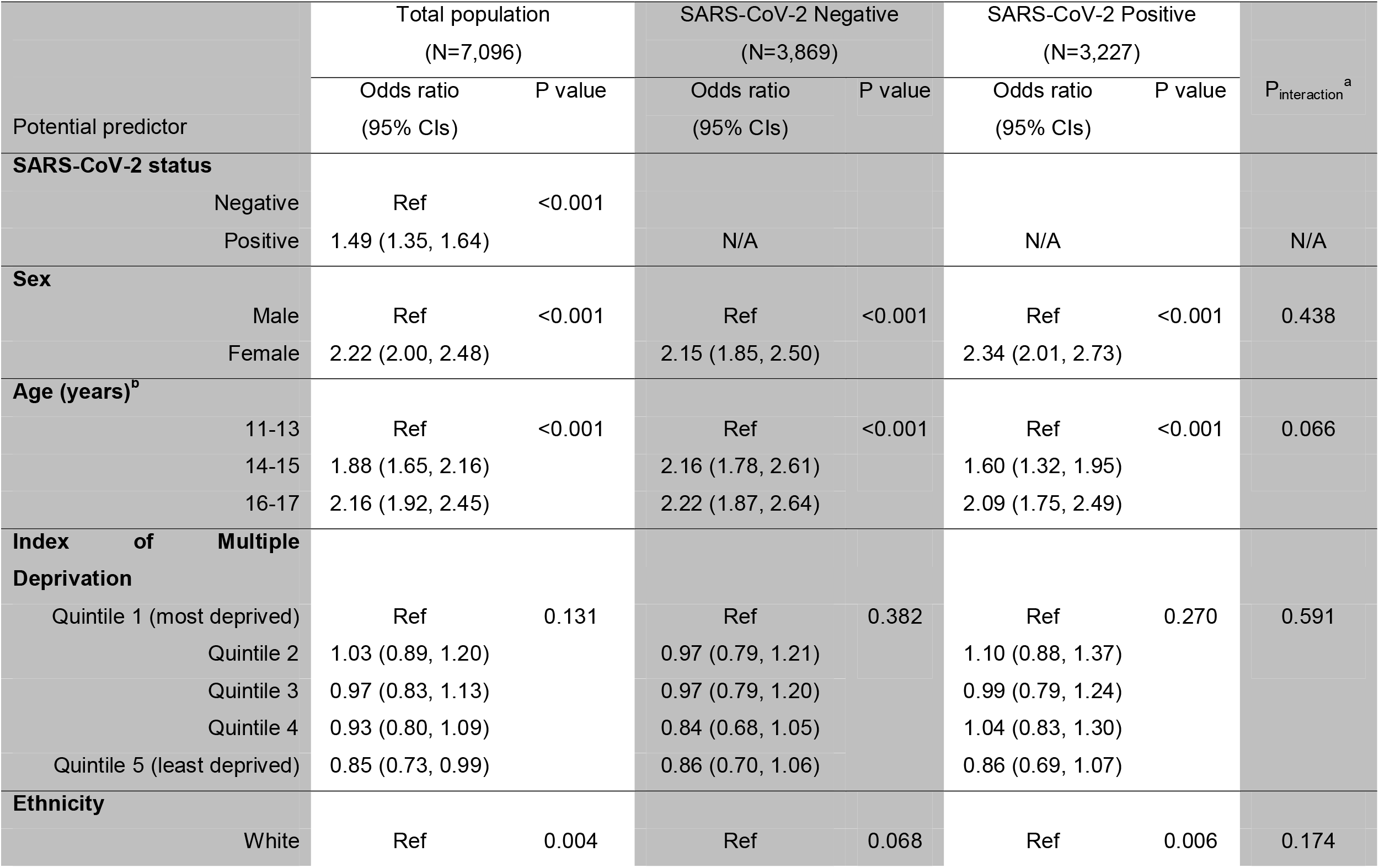
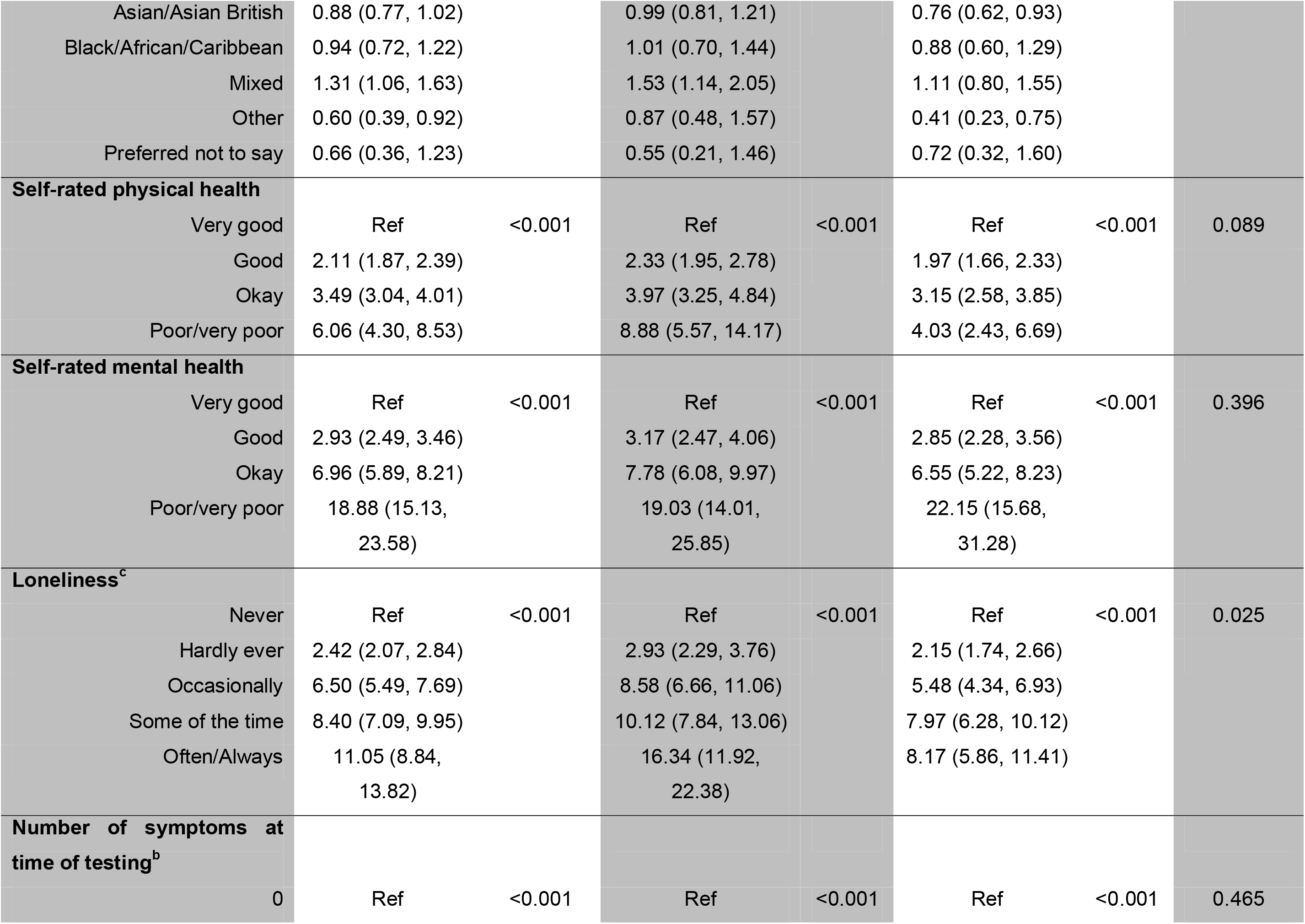
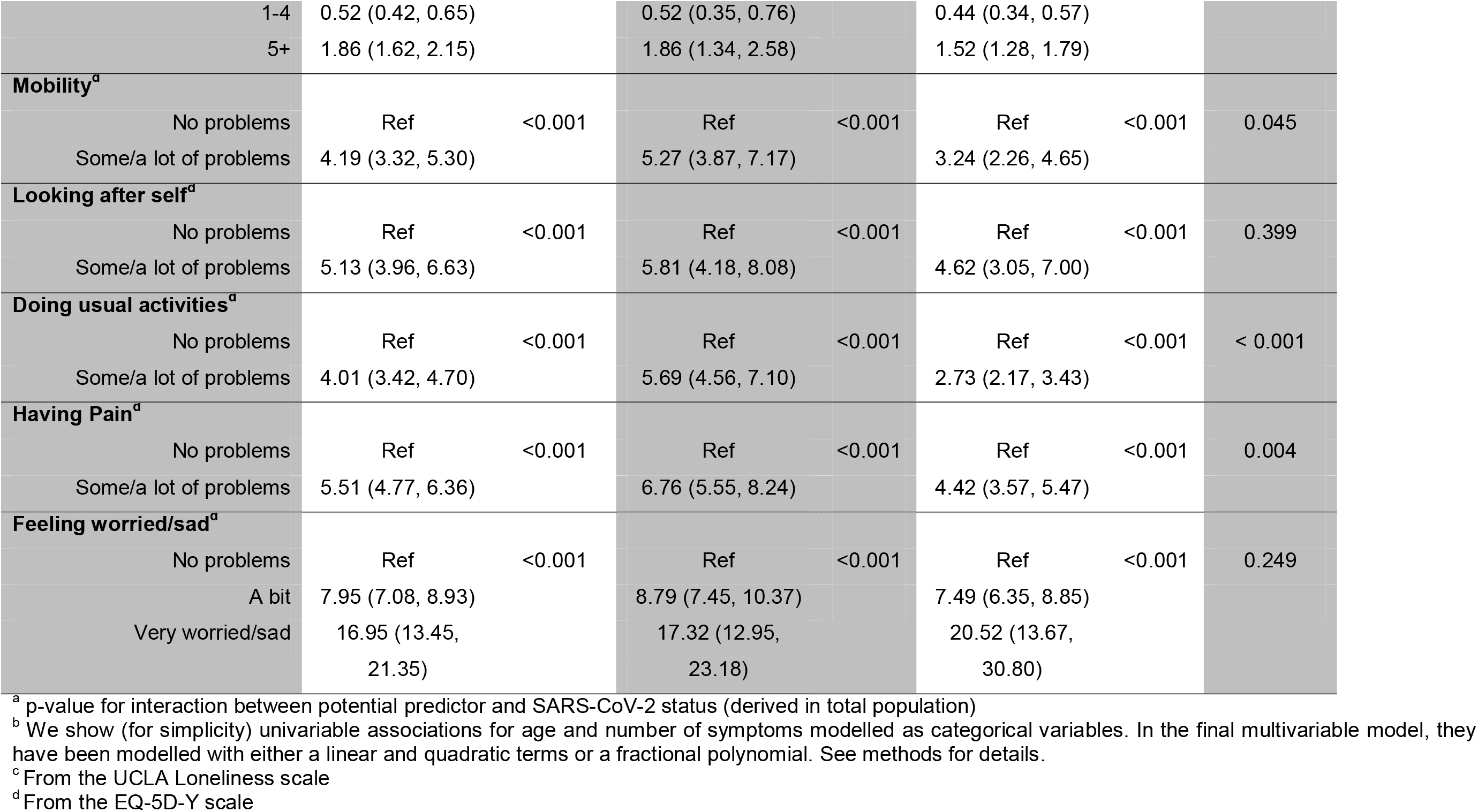
Odds ratios (95% CIs) of univariable associations between potential predictors and at least one persistent impairing physical symptom 3 months after a PCR-test, overall and stratified by SARS-CoV-2 status

### Multivariable predictive model

In the final developed model (eTable 2), SARS-COV-2 status, number of symptoms at testing, sex, age, ethnicity, self-rated physical and mental health, feelings of loneliness and four items from the EQ-5D-Y scale (problems looking after self, doing usual activities, having pain, feeling worried/sad) before testing predicted the outcome. The impact of some predictors differed by SARS-COV-2 status: interactions between SARS-COV-2 status and age, ethnicity, self-rated mental health, feelings of loneliness, and problems doing usual activities were retained. The supplementary figures show graphs from the final developed model, for all included predictors, of the probability of having the outcome.

### Model Performance

The model showed excellent calibration and discrimination. It was perfectly calibrated in the model development data with an apparent slope of 1 and an apparent calibration-in-the-large of 0 (eTable 3). Good overall model calibration was further confirmed by the calibration plot (Figure 1), with narrow confidence intervals and closely aligned predicted and observed probabilities for 10 equally sized risk groups. The predictive model showed strong discrimination with a C-statistic of 0.841(95% CI:0.831,0.850). Bootstrap internal validation showed small model overfitting with an optimism corrected C slope close to one. The bootstrapping approach provided a shrinkage factor of 0.97527. We also generated the heuristic shrinkage factor (again close to one: 0.98399). We chose the bootstrap shrinkage factor as it was slightly smaller, and, applied it to the original β coefficients to obtain the optimism adjusted coefficients before re-estimating the intercept for the final model given in Box 1 (Supplementary material) and eTable 2.

### Worked examples

Box 1 (Supplementary material) shows the prediction equation for estimating the risk of experiencing at least one impairing physical symptom 3 months post-PCR-test in 11-to-17-year-old CYP. We demonstrate with hypothetical examples the predicted risk of impairing physical symptom 3 months post-test in Table 4. A calculator is provided in the Supplementary section.

**Table 4:** Hypothetical examples of predicted risk of persistent impairing physical symptom 3 months after a PCR-test, using our prediction model

| Characteristic | Examples |  |  |  |  |  |
| --- | --- | --- | --- | --- | --- | --- |
|  | 1 | 2 | 3 | 4 | 5 | 6 |
| SARS-CoV-2 status | Positive | Negative | Positive | Negative | Positive | Negative |
| Sex | Female | Female | Male | Male | Female | Female |
| Age (years) | 17 | 17 | 13 | 13 | 17 | 17 |
| Ethnicity | White | White | White | White | White | White |
| Self-rated physical health | Okay | Okay | Okay | Okay | Good | Good |
| Self-rated mental health | Okay | Okay | Okay | Okay | Good | Good |
| Loneliness | Some of the time | Some of the time | Some of the time | Some of the time | Hardly Ever | Hardly Ever |
| Number of symptoms at time of testing | 0 | 0 | 0 | 0 | 3 | 3 |
| Problems looking after myself | No problems | No problems | No problems | No problems | No problems | No problems |
| Problems doing usual activities | No problems | No problems | No problems | No problems | No problems | No problems |
| Having pain | No problems | No problems | No problems | No problems | No problems | No problems |
| Feeling worried/sad** | A bit worried | A bit worried | A bit worried | A bit worried | No problems | No problems |
| Predicted Risk | 0.767 | 0.570 | 0.667 | 0.503 | 0.167 | 0.083 |

For details on how the predicted risk was calculated see supplementary section for formula (Box 1) and calculator.

As an example, the predicted risk of outcome for a 14-year-old, white male, with no symptoms at testing, very good physical health, never feeling lonely, no problems on all included EQ-5D-Y items and poor/very poor mental health before testing would be 0.19 if he tested positive and 0.06 if he tested negative; if he had very good mental health before testing, the risk would be 0.08 if positive and 0.04 if negative.

## Discussion

To our knowledge, we have developed the first risk prediction model that uses self-reported information from CYP to estimate their probability of experiencing at least one impairing physical symptom 3 months after SARS-COV-2 testing. SARS-COV-2 status, number of physical symptoms at testing, sex, age, ethnicity, self-rated physical and mental health, feelings of loneliness and four items from the EQ-5D-Y scale (all before testing) predicted having at least one impairing physical symptom 3 months later, with the impact of some predictors differing by SARS-COV-2 status. We provide a risk calculator to predict CYP most likely to experience impairing physical symptoms, to triage those who need support and for whom early intervention might be of greatest benefit. Importantly, our model has excellent predictive ability, calibration and discrimination. It enables us to answer important clinical questions such as ‘are those who have many symptoms during acute SARS-COV-2 infection at greater risk of ‘Long COVID’ than those without?’. The answer is ‘yes’ but our model provides a more nuanced answer by considering other factors.

Our goal was to provide a model that utilizes multiple factors (i.e., predictors) in combination, to accurately predict experiencing at least one impairing physical symptom 3 months post-test. Importantly, our focus was not on whether included predictors are causal or not. Instead, the focus was overall predictive performance of the model.^32^ As such, we followed the guidelines to model building.^26^ The large sample allowed flexible examination of the potential for relationships to differ by SARS-COV-2 status and by the shape of the association without considerable concerns about overfitting. Model fitting statistics were extremely favorable and the use of a matched national cohort sample of test-positive and test-negative CYP is unique. As this is the first study of its kind, results need to be externally validated on other independent datasets and in different populations and settings. Additionally, the model will be reassessed for experiencing at least one impairing physical symptom beyond 3 months using CLoCk data at 6, 12 and 24 months after testing as data become available. It is possible many of the predictors stay the same but acknowledge there may be differences as the disease profile (and, therefore, predictors) change over the course of the illness.

We acknowledge study limitations. Baseline measures (at/or before testing) were subject to recall bias because they were not taken at the time of acute infection. We were unable to assess whether symptoms waxed and waned between testing and questionnaire. The CLoCk study response rate (13.4%)^4^ is typical of surveys of this type;^33^ additionally, our participants are largely representative of the target population as a whole.^4^ Nevertheless, the possibility of selection bias in both directions (CYP more likely to participate if they have persistent symptoms, or less likely to participate if too unwell) among respondents cannot be ruled out. Furthermore, as the background epidemiological situation in relation to SARS-COV-2 infection prevalence changes, there is a need to reassess possible differences in our model’s predictive value over time. Finally, caution is required for predictions based on data extrapolation/situations where there are only a very small number of observations for different predictor combinations.

To our knowledge, no other study has explicitly aimed to develop a risk prediction model for experiencing impairing physical symptoms several months after SARS-COV-2 testing. Moreover, the majority of previous studies lack a SARS-CoV-2 test-negative comparison group and so distinguishing long-term symptoms predicted by SARS-CoV-2 infection from background rates or pandemic-related effects remains a challenge.^5^ More recent studies include control groups and, thus, broad comparisons can be made. Our finding that the odds of experiencing at least one impairing physical symptom 3 months post-test was 1.49 times higher in SARS-CoV-2 positive compared to SARS-CoV-2 negative CYP, is in line with findings from the LongCOVIDKidsDK study, where the SARS-CoV-2 test-positive group had 1.22 times higher odds of having at least one ‘Long COVID’ symptom lasting at least 2 months compared with the SARS-CoV-2 test-negative group.^34^ We found both test-positive and test-negative CYP had impairing physical symptoms 3 months post-test with a difference of 9.3% between these groups. In contrast, in a Danish study, the prevalence of reported symptoms in CYP aged 6-17 years lasting more than 4 weeks was similar regardless of SARS-CoV-2 status (28% test-positives; 27.2% test-negatives).^18^ Discrepancies in findings could be due to several reasons including timing of outcome (>4 weeks vs ∼3 months) and differences in recruitment methodology, recruitment rates between test-positives and test-negatives or underlying prevalence levels in the countries at the time of study. Our results are consistent with findings in adults, where number of symptoms at onset^35^ and female sex^36^ were associated with ‘Long COVID’, and pre-existing diagnosis of depression/anxiety being over-represented in those with fatigue after SARS-CoV-2 infection.^36^.

Understanding which CYP are at risk of experiencing impairing physical symptoms is important for individuals (e.g., in decision making about whether to receive COVID-19 vaccination) and health services provision (e.g., for careful monitoring, early intervention, and hopefully reduction in the burden of prolonged health problems). In conclusion, using data from a large national matched cohort study, we developed a prediction model for experiencing at least one impairing physical symptom 3 months after SARS-COV-2 testing in CYP. Our model has excellent performance, and we hope it will serve as a useful tool for early identification and management of CYP at risk of persisting physical symptoms in the context of the COVID-19 pandemic.

## Supporting information

Risk calculator

disclosure form

## Data Availability

All data produced in the present work are contained in the manuscript

## Acknowledgement

Michael Lattimore, Public Health England, as Project Officer for the CLoCk study

## Conflicts of interest

Terence Stephenson is Chair of the Health Research Authority and therefore recused himself from the research ethics application. All other authors declare no competing interests.

The study was approved by Yorkshire and the Humber–South Yorkshire Research Ethics Committee (reference: 21/YH/0060).

TS, RS, SMPP, BLS, MDN conceived the study. MDN conducted the statistical analyses, accessed, and verified the data and drafted the manuscript. Both SMPP and BLS provided statistical input to the design, accessed and verified the data and drafted the manuscript. TS and RS supported the drafting of the manuscript. All authors (MDN, TS, RS, BLS, SNL, RS, KM, NR, EYC, TF, IH, EC, SMPP) critically revised the manuscript for important intellectual content and gave final approval for the version to be published. All authors agree to be accountable for all aspects of the work in ensuring that questions related to the accuracy or integrity of any part of the work are appropriately investigated and resolved.

## Funding statement

Funded by The Department of Health and Social Care, in their capacity as the National Institute for Health Research (NIHR), and by UK Research & Innovation (UKRI) who have awarded funding grant number COVLT0022. All research at Great Ormond Street Hospital NHS Foundation Trust and UCL Great Ormond Street Institute of Child Health is made possible by the NIHR Great Ormond Street Hospital Biomedical Research Centre. The views expressed are those of the author(s) and not necessarily those of the NHS, the NIHR or the Department of Health. SMPP is supported by a UK Medical Research Council Career Development Award (reference: MR/P020372/1).

## Notes

### Author Declarations

We use data from the CLoCk study: a national cohort study of SARS-CoV-2 PCR-positive CYP aged 11-17 years living in England who were matched at study invitation, on month of test, age, sex, and geographical area, to SARS-CoV-2 test-negative CYP selected from the national testing database at Public Health England (now UKHSA).20 The study was approved by Yorkshire and the Humber South Yorkshire Research Ethics Committee (reference: 21/YH/0060). 20.Stephenson T, Shafran R, De Stavola B, et al. Long COVID and the mental and physical health of children and young people: national matched cohort study protocol (the CLoCk study). BMJ Open. Aug 26 2021;11(8):e052838. doi:10.1136/bmjopen-2021-052838

