## Supplementary material for "Developing a model for predicting impairing physical symptoms in children 3 months after a SARS-CoV-2 PCR-test: The CLoCk Study": Figure 1

**Figure 1.** Observed and predicted risk of persistent impairing physical symptoms 3 months after a PCR-test^a^

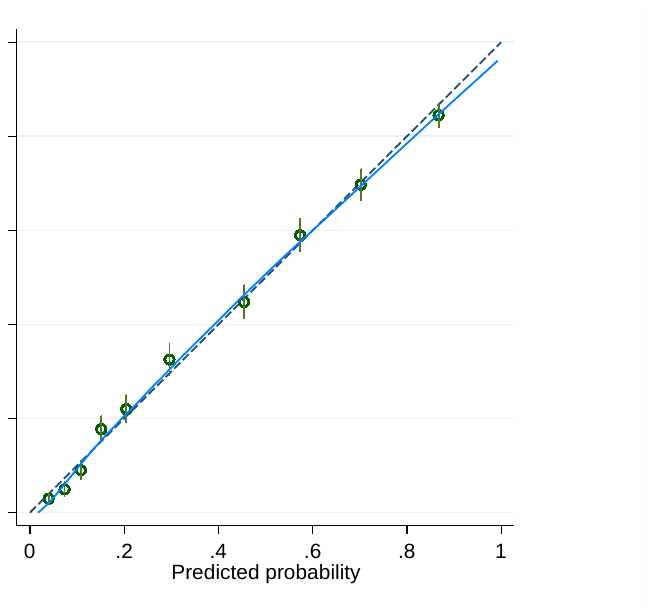

^a^ This graph shows the mean predicted probability (hollow dots) and 95% confidence intervals of persistent impairing physical symptoms 3 months after a PCR-test plotted against the observed proportion of the same outcome for 10 equally sized groups. The dashed line represents the line of equality and perfect calibration. The blue solid line is a smoothed locally weighted scatter plot smoothing (Lowess) regression line.
