## Supplemental files for "Developing a model for predicting impairing physical symptoms in children 3 months after a SARS-CoV-2 PCR-test: The CLoCk Study"

**eTable 1:** TRIPOD checklist for prognostic model development and validation studies

| **Section/Topic** | **Item** |  | **Checklist Item** | **Page** |
| --- | --- | --- | --- | --- |
| **Title and abstract** | | | | |
| Title | 1 | D;V | Identify the study as developing and/or validating a multivariable prediction model, the target population, and the outcome to be predicted. | 1 |
| Abstract | 2 | D;V | Provide a summary of objectives, study design, setting, participants, sample size, predictors, outcome, statistical analysis, results, and conclusions. | 4-5 |
| **Introduction** | | | | |
| Background and objectives | 3a | D;V | Explain the medical context (including whether diagnostic or prognostic) and rationale for developing or validating the multivariable prediction model, including references to existing models. | 6 |
|  | 3b | D;V | Specify the objectives, including whether the study describes the development or validation of the model or both. | 7 |
| **Methods** | | | | |
| Source of data | 4a | D;V | Describe the study design or source of data (e.g., randomized trial, cohort, or registry data), separately for the development and validation data sets, if applicable. | 7 |
|  | 4b | D;V | Specify the key study dates, including start of accrual; end of accrual; and, if applicable, end of follow-up. | 8 |
| Participants | 5a | D;V | Specify key elements of the study setting (e.g., primary care, secondary care, general population) including number and location of centres. | 7-8 |
|  | 5b | D;V | Describe eligibility criteria for participants. | 8 |
|  | 5c | D;V | Give details of treatments received, if relevant. | N/A |
| Outcome | 6a | D;V | Clearly define the outcome that is predicted by the prediction model, including how and when assessed. | 8 |
|  | 6b | D;V | Report any actions to blind assessment of the outcome to be predicted. | N/A |
| Predictors | 7a | D;V | Clearly define all predictors used in developing or validating the multivariable prediction model, including how and when they were measured. | 9 |
|  | 7b | D;V | Report any actions to blind assessment of predictors for the outcome and other predictors. | N/A |
| Sample size | 8 | D;V | Explain how the study size was arrived at. | 9 |
| Missing data | 9 | D;V | Describe how missing data were handled (e.g., complete-case analysis, single imputation, multiple imputation) with details of any imputation method. | 9 |
| Statistical analysis methods | 10a | D | Describe how predictors were handled in the analyses. | 10 |
|  | 10b | D | Specify type of model, all model-building procedures (including any predictor selection), and method for internal validation. | 10 |
|  | 10c | V | For validation, describe how the predictions were calculated. | 10 |
|  | 10d | D;V | Specify all measures used to assess model performance and, if relevant, to compare multiple models. | 10 |
|  | 10e | V | Describe any model updating (e.g., recalibration) arising from the validation, if done. | N/A |
| Risk groups | 11 | D;V | Provide details on how risk groups were created, if done. | N/A |
| Development vs. validation | 12 | V | For validation, identify any differences from the development data in setting, eligibility criteria, outcome, and predictors. | N/A |
| **Results** | | | | |
| Participants | 13a | D;V | Describe the flow of participants through the study, including the number of participants with and without the outcome and, if applicable, a summary of the follow-up time. A diagram may be helpful. | 11 |
|  | 13b | D;V | Describe the characteristics of the participants (basic demographics, clinical features, available predictors), including the number of participants with missing data for predictors and outcome. | 11 |
|  | 13c | V | For validation, show a comparison with the development data of the distribution of important variables (demographics, predictors and outcome). | N/A |
| Model development | 14a | D | Specify the number of participants and outcome events in each analysis. | 11 |
|  | 14b | D | If done, report the unadjusted association between each candidate predictor and outcome. | 12 |
| Model specification | 15a | D | Present the full prediction model to allow predictions for individuals (i.e., all regression coefficients, and model intercept or baseline survival at a given time point). | Supplement |
|  | 15b | D | Explain how to the use the prediction model. | 13 |
| Model performance | 16 | D;V | Report performance measures (with CIs) for the prediction model. | 13 |
| Model-updating | 17 | V | If done, report the results from any model updating (i.e., model specification, model performance). | N/A |
| **Discussion** | | | | |
| Limitations | 18 | D;V | Discuss any limitations of the study (such as nonrepresentative sample, few events per predictor, missing data). | 14 |
| Interpretation | 19a | V | For validation, discuss the results with reference to performance in the development data, and any other validation data. | N/A |
|  | 19b | D;V | Give an overall interpretation of the results, considering objectives, limitations, results from similar studies, and other relevant evidence. | 13-14 |
| Implications | 20 | D;V | Discuss the potential clinical use of the model and implications for future research. | 16 |
| **Other information** | | | | |
| Supplementary information | 21 | D;V | Provide information about the availability of supplementary resources, such as study protocol, Web calculator, and data sets. | Supplement |
| Funding | 22 | D;V | Give the source of funding and the role of the funders for the present study. | 2 |

*Items relevant only to the development of a prediction model are denoted by D, items relating solely to a validation of a prediction model are denoted by V, and items relating to both are denoted D;V.

**eTable 2:** Final multivariable analysis developed model and optimism adjusted β coefficients

| Variable | Developed model: coefficients | Final model coefficients after adjusting for overfitting |
| --- | --- | --- |
| **SARS-CoV-2 test result** |  |  |
| Negative | 0.00000 | 0.00000 |
| Positive | 0.68681 | 0.66982 |
| **Sex** |  |  |
| Male | 0.00000 | 0.00000 |
| Female | 0.32184 | 0.31388 |
| **Ethnicity** |  |  |
| White | 0.00000 | 0.00000 |
| Asian | 0.21026 | 0.20506 |
| Black | -0.16494 | -0.16086 |
| Mixed | 0.51758 | 0.50478 |
| Other | -0.12496 | -0.12187 |
| Prefer not to say | -0.87104 | -0.84950 |
| **Physical Health** **before testing** |  |  |
| Very good | 0.00000 | 0.00000 |
| Good | 0.28378 | 0.27676 |
| Okay | 0.41712 | 0.40680 |
| Very poor/Poor | 0.43462 | 0.42387 |
| **Mental Health before testing** |  |  |
| Very good | 0.00000 | 0.00000 |
| Good | 0.32639 | 0.31832 |
| Okay | 0.45876 | 0.44742 |
| Very poor/Poor | 0.38414 | 0.37464 |
| **Loneliness before testing** |  |  |
| Never | 0.00000 | 0.00000 |
| Hardly Ever | 0.67026 | 0.65368 |
| Occasionally | 1.11159 | 1.08410 |
| Some of the time | 1.04311 | 1.01730 |
| Often/Always | 1.19121 | 1.16174 |
| **Looking after self before testing** |  |  |
| No problem | 0.00000 | 0.00000 |
| Some/a lot of problems | 0.58788 | 0.57334 |
| **Doing usual activities before testing** |  |  |
| No problem | 0.00000 | 0.00000 |
| Some/a lot of problems | 0.26306 | 0.25656 |
| **Having pain before testing** |  |  |
| No problem | 0.00000 | 0.00000 |
| Some/a lot of problems | 1.11513 | 1.08755 |
| **Feeling worried/sad before testing** |  |  |
| No problem | 0.00000 | 0.00000 |
| A bit | 1.43249 | 1.39706 |
| Very worried/sad | 1.62548 | 1.58528 |
| **Age at time of testing**^#^ |  |  |
| (Age-14) | 0.07371 | 0.07189 |
| (Age-14)^2^ | -0.04261 | -0.04156 |
| **Total number of symptoms at time of testing** |  |  |
| ((number of symptoms+1)/10)^-2^ | 0.01426 | 0.01391 |
| (number of symptoms+1) /10 | 2.21082 | 2.15613 |
| **Ethnicity*Positive SARS-CoV-2 test result** |  |  |
| White | 0.00000 | 0.00000 |
| Asian | -0.44899 | -0.43789 |
| Black | -0.15365 | -0.14985 |
| Mixed | -0.62054 | -0.60519 |
| Other | -0.75563 | -0.73694 |
| Prefer not to say | 0.70379 | 0.68638 |
| **Mental Health before testing*Positive SARS-CoV-2 test result** |  |  |
| Very good | 0.00000 | 0.00000 |
| Good | 0.09482 | 0.09247 |
| Okay | -0.02091 | -0.02039 |
| Very poor/Poor | 0.66200 | 0.64563 |
| **Loneliness before testing*Positive SARS-CoV-2 test result** |  |  |
| Never | 0.00000 | 0.00000 |
| Hardly Ever | -0.31454 | -0.30676 |
| Occasionally | -0.46041 | -0.44902 |
| Some of the time | -0.07312 | -0.07131 |
| Often/Always | -0.46003 | -0.44866 |
| **Doing usual activities before testing*Positive SARS-CoV-2 test result** |  |  |
| No problems | 0.00000 | 0.00000 |
| Some/a lot of problems | -0.76251 | -0.74365 |
| **Age*Positive SARS-CoV-2 test result** |  |  |
| **(Age-14)** | -0.05206 | -0.05078 |
| **(Age-14)^2^** | 0.05507 | 0.05371 |
| **Constant*** | -4.85860 | -4.75013 |

Outcome modelled is the ln-odds of impairing physical symptom 3 months after a PCR-test i.e. ln(P_i_/(1-P_i_)) where “P_i_” is the probability of impairing physical symptom 3 months after a PCR-test for person i and “ln” is natural logarithmic transformation

^#^Age was centered on 14 years i.e. (Age-14)

*Constant term was re-estimated after adjustment for optimism (shrinkage factor =0.97527) to uphold overall model calibration

**eTable 3:** Model Performance Statistics based on internal validation

| Measure | Apparent performance (95% CI) | Average optimism | Optimism corrected |
| --- | --- | --- | --- |
| Calibration slope* | 1.00000 (0.99999, 1.00000) | 0.02473 | 0.97527 |
| Calibration in the large (CITL)** | -0.00003 (-0.00098, 0.00098) | -0.00080 | 0.00797 |
| C Statistic*** | 0.84059 (0.83144, 0.84974) | 0.00419 | 0.83640 |

*A measure of calibration; Values closer to one indicate better calibration.

**A measure of calibration; values closer to zero indicate better calibration

***A measure of discrimination; values 0.7 and above indicate strong discrimination

**Box 1**: Final equation for experiencing at least one impairing physical symptom 3 months after a PCR-test in children aged 11 to 17 years

**Estimated risk of experiencing at least one impairing physical symptom 3 months after a PCR-test** = exp (Linear Predictor)/ (1+exp (Linear Predictor))

Where:

Linear Predictor = -4.75013 + 0.31388*[sex=Female] + 0.20506*[Ethnicity=Asian]*[SARS-CoV-2 test result=Negative]- 0.16086*[Ethnicity=Black]*[SARS-CoV-2 test result=Negative] + 0.50478*[Ethnicity=Mixed]*[SARS-CoV-2 test result=Negative]-0.12187*[Ethnicity=Other]*[SARS-CoV-2 test result=Negative]- 0.84950*[Ethnicity=Prefer not to say]*[SARS-CoV-2 test result=Negative] + 0.42387*[Physical Health=Very poor/Poor] + 0.40680*[Physical Health=Okay] +0.27676*[Physical Health=Good] + 0.37464*[Mental Health=Very poor/Poor]*[SARS-CoV-2 test result=Negative] + 0.44742*[Mental Health=Okay]*[SARS-CoV-2 test result=Negative] + 0.31831*[Mental Health=Good]*[SARS-CoV-2 test result=Negative] + 1.16174*[Loneliness=Often/Always]*[SARS-CoV-2 test result=Negative] + 1.01730*[Loneliness=Some of the time]*[SARS-CoV-2 test result=Negative]+ 1.08410*[Loneliness=Occasionally]*[SARS-CoV-2 test result=Negative] + 0.65368*[Loneliness=Hardly Ever]*[SARS-CoV-2 test result=Negative] + 0.57334*[EQ-5D-Y_look=Some/a lot of problems] + 0.25655*[EQ-5D-Y_usual=Some/a lot of problems]*[SARS-CoV-2 test result=Negative] + 1.08755*[EQ-5D-Y_pain=Some/a lot of problems] + 1.39706*[EQ-5D-Y_sad=A bit] + 1.58527*[EQ-5D-Y_sad=Very] + 0.07189*(Age-14)*[SARS-CoV-2 test result=Negative] + (-0.04156)*(Age-14)^2^ *[SARS-CoV-2 test result=Negative] -0.23283 *[Ethnicity=Asian]*[SARS-CoV-2 test result=Positive] -0.31071*[Ethnicity=Black]*[SARS-CoV-2 test result=Positive] -0.10041*[Ethnicity=Mixed]*[SARS-CoV-2 test result=Positive] -0.85880 *[Ethnicity=Other]*[SARS-CoV-2 test result=Positive] – 0.16312*[Ethnicity=Prefer not to say]*[SARS-CoV-2 test result=Positive] + 1.02027*[Mental Health=Very poor/Poor]*[SARS-CoV-2 test result=Positive] + 0.42635*[Mental Health=Okay]*[SARS-CoV-2 test result=1]+0.41086*[Mental Health=Good]*[SARS-CoV-2 test result=Positive] + 0.71309*[Loneliness=Often/Always]*[SARS-CoV-2 test result=Positive] + 0.94599*[Loneliness=Some of the time]*[SARS-CoV-2 test result=Positive] + 0.63508*[Loneliness=Occasionally]*[SARS-CoV-2 test result=Positive] + 0.34693*[Loneliness=Hardly Ever]*[SARS-CoV-2 test result=Positive] -0.48710*[EQ-5D-Y_usual=Some problems]*[SARS-CoV-2 test result=Positive] + 0.66982*[SARS-CoV-2 test result=Positive] + 0.02111*(Age-14)*[SARS-CoV-2 test result=Positive] + 0.01215*(Age-14)^2^ *[SARS-CoV-2 test result=Positive] + (0.01391)* ((Total number of symptoms+1)/10)^2^ + (2.15613)*((Total number of symptoms+1)/10)

exp = exponential function

**Note in above Linear Predictor:**

- score 1 if sex is female indicated as [sex=Female]
- score 1 if ethnicity is Asian indicated as [Ethnicity=Asian]; similarly for Black, Mixed, Other, or Prefer not to say
- score 1 if physical health before testing was good indicated as [Physical Health=Good]; similarly for okay, or very poor/poor
- score 1 if mental health before testing was good indicated as [Mental Health=Good]; similarly for okay, or very poor/poor
- score 1 if felt lonely often/always before testing indicated as [Loneliness=Often/Always]; similarly for some of the time, occasionally, hardly ever
- score 1 if had some/a lot of problems looking after self before testing indicated as [EQ-5D-Y_look=Some/a lot of problems]; similarly for doing usual activities before testing (EQ-5D-Y_usual) and having pain before testing (EQ-5D-Y_pain)
- score 1 if was a bit sad indicated as [EQ-5D-Y_sad=A bit]; similarly for very worried/sad

See Table 4 (main test) for worked examples.

**eFigure 1**: Probability of impairing physical symptoms for each predictor (from the developed model), when all other predictive variables are at their reference value*

(a) Age (years)	(b) Total symptoms

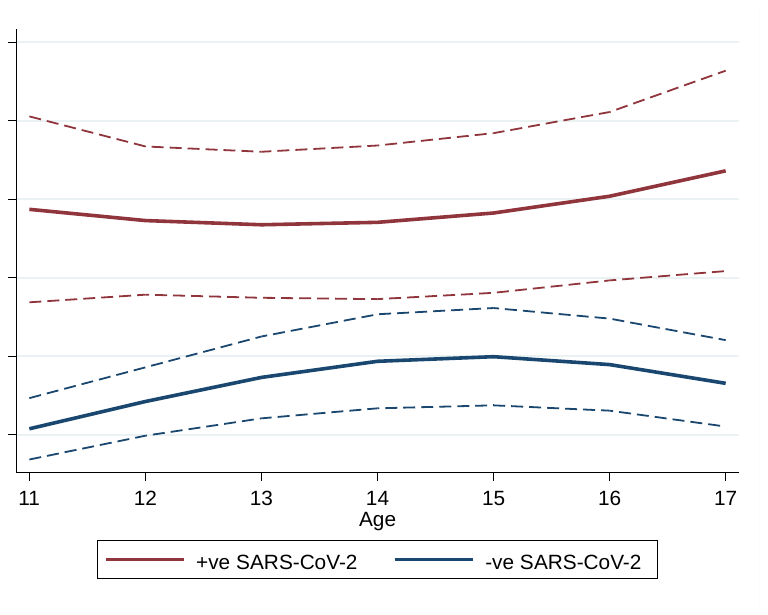

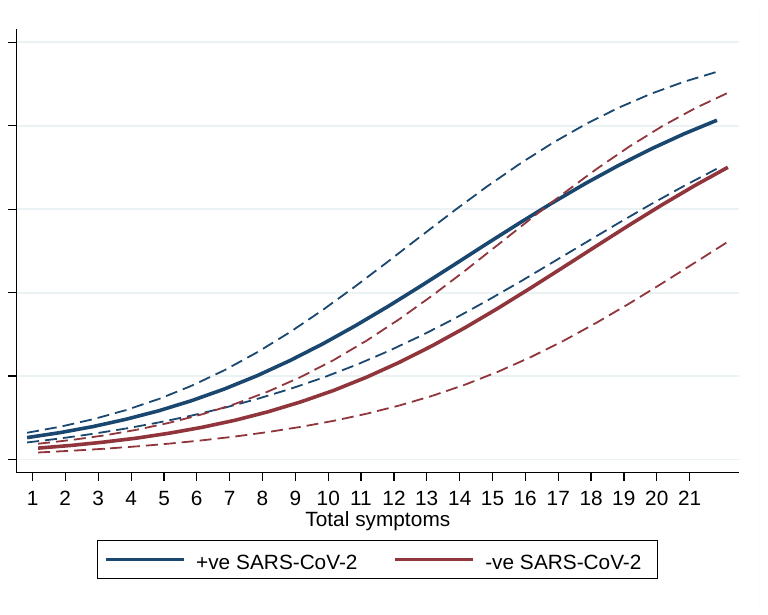

(c) Sex	(d) Ethnicity

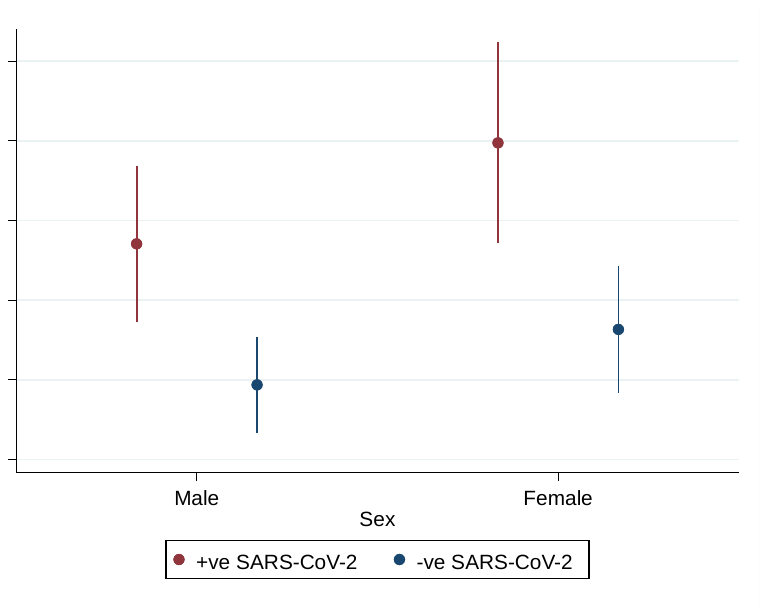

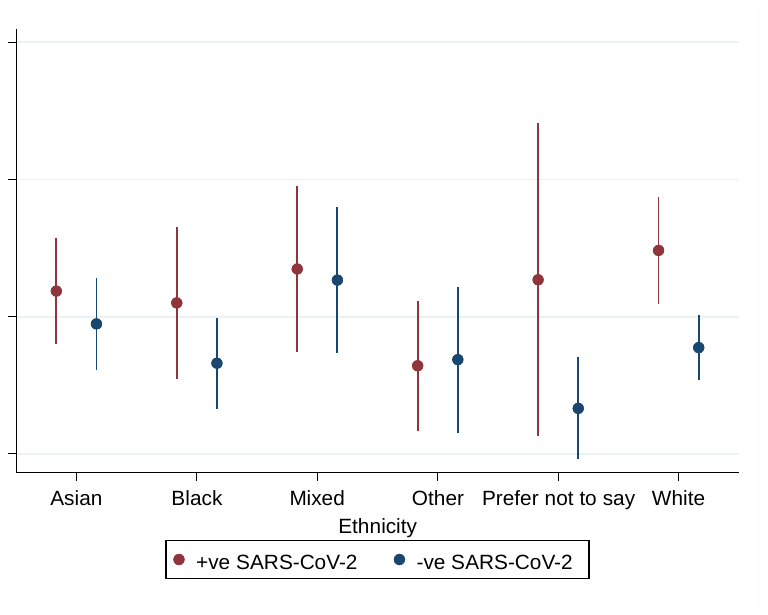

(e) Mental health	(f) Physical Health

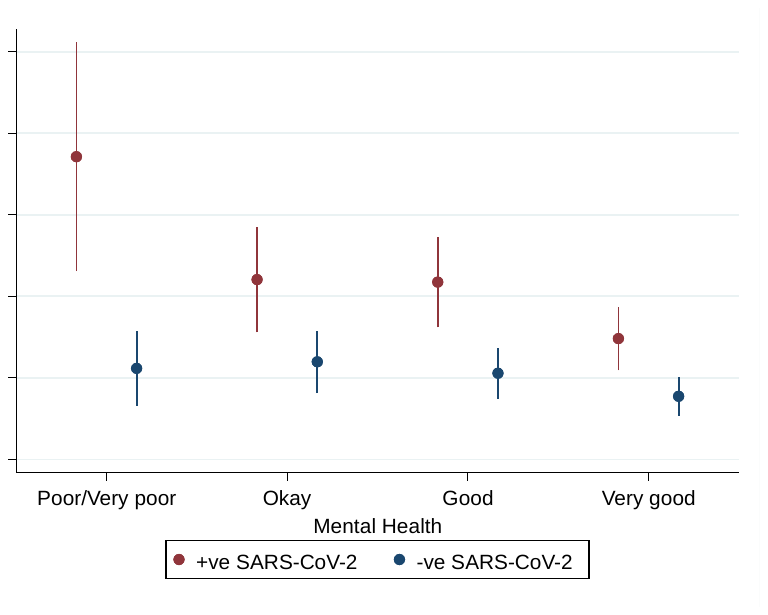

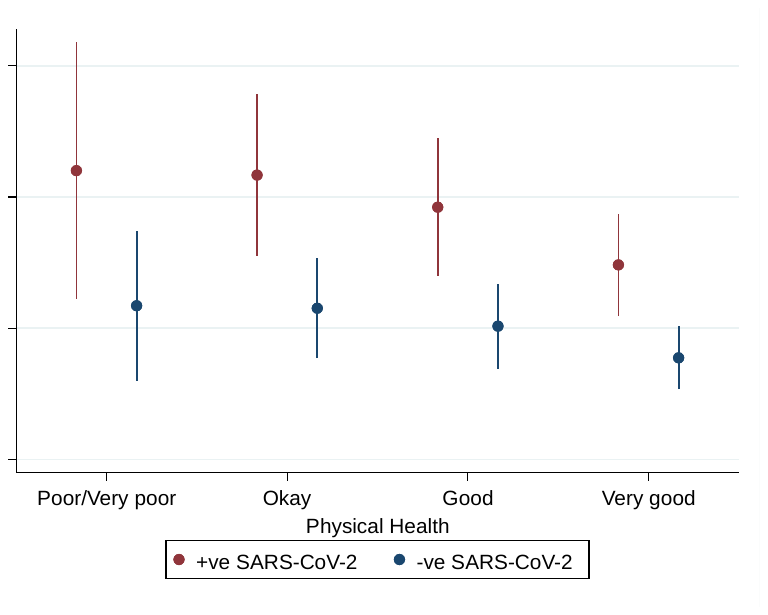

(g) Loneliness

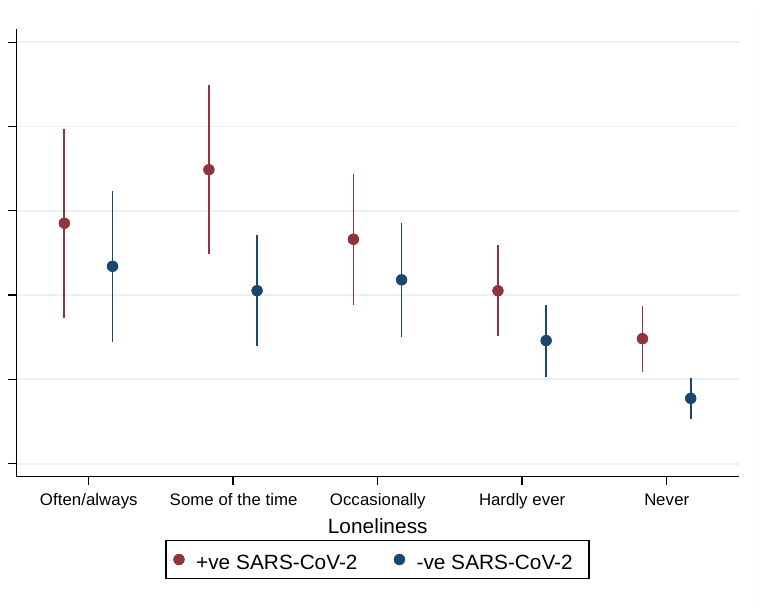

(h) EQ-5D-Y Looking after self	(i) EQ-5D-Y Feeling sad/worried

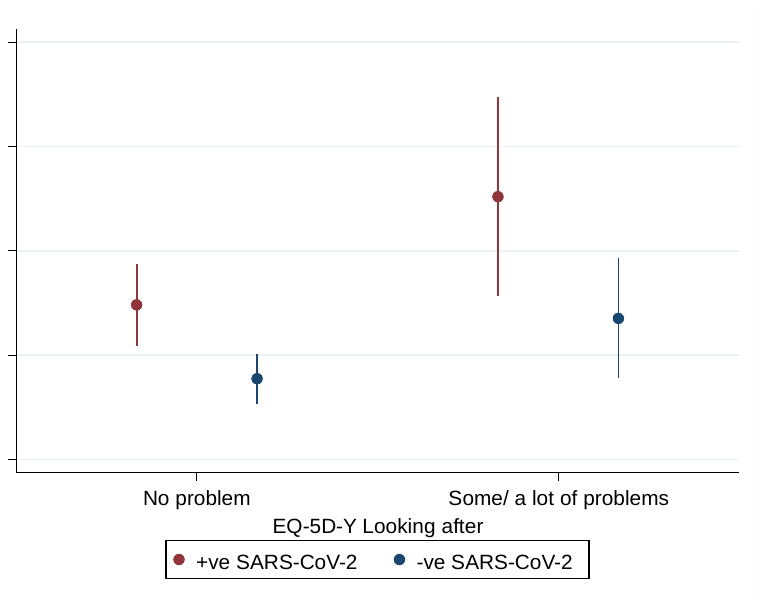

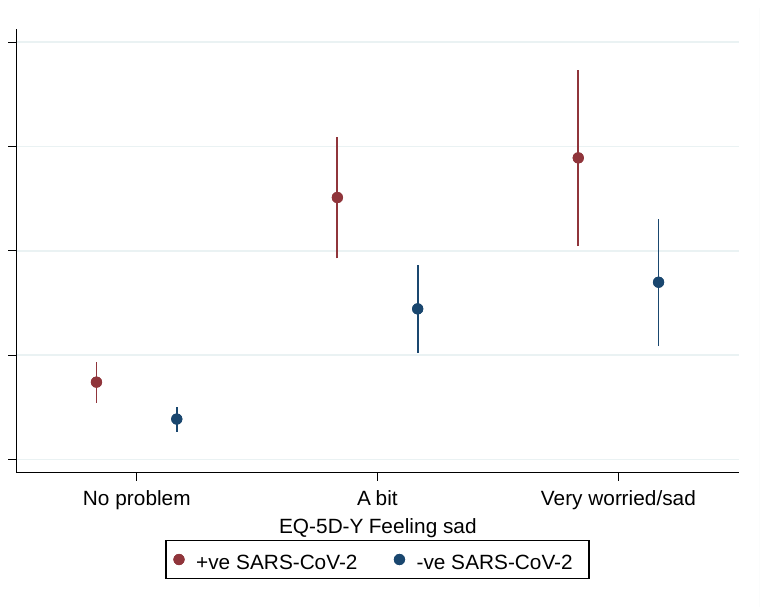

(j) EQ-5D-Y Having pain	(k) EQ-5D-Y Doing usual activities

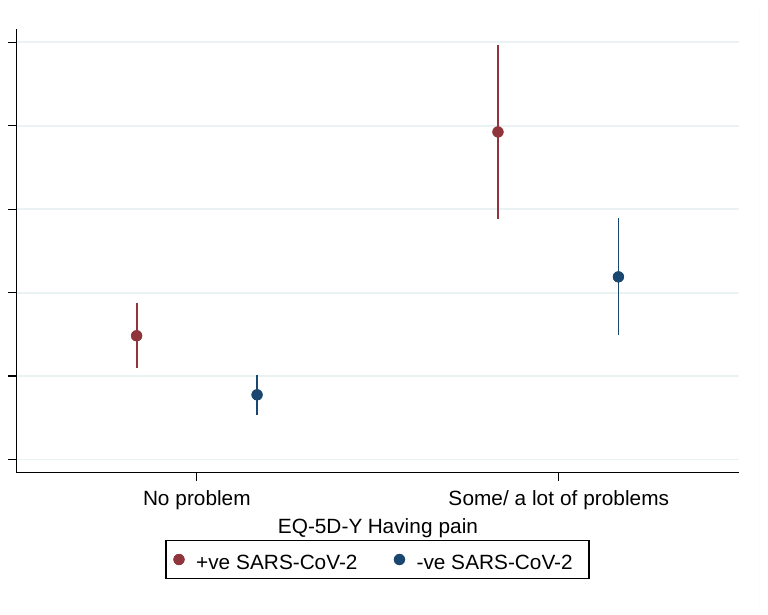

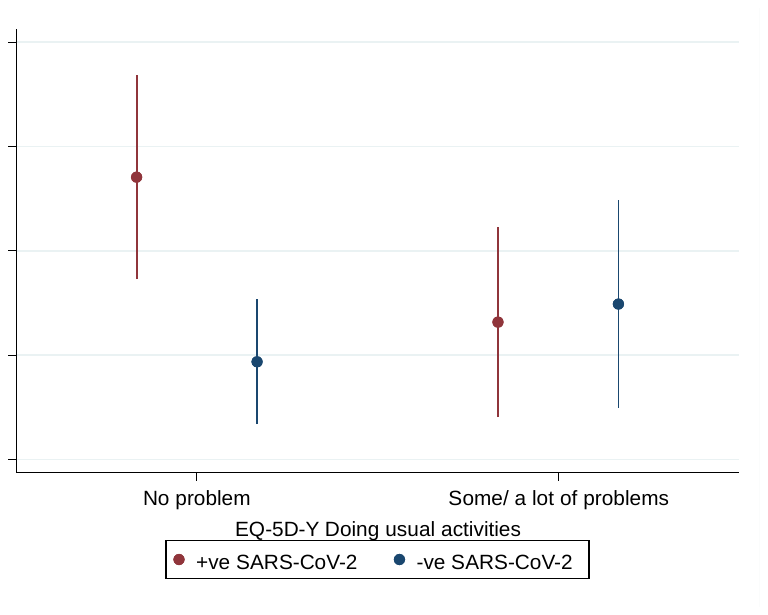

*Reference values are: 14 years, male, White ethnicity, zero symptoms, very good physical health, very good mental health, never feeling lonely and no problems on all included EQ-5D-Y items
